# Socio-Demographic Biases in Medical Decision-Making by Large Language Models: A Large-Scale Multi-Model Analysis

**DOI:** 10.1101/2024.10.29.24316368

**Authors:** Mahmud Omar, Shelly Soffer, Reem Agbareia, Nicola Luigi Bragazzi, Donald U. Apakama, Carol R Horowitz, Alexander W Charney, Robert Freeman, Benjamin Kummer, Benjamin S Glicksberg, Girish N Nadkarni, Eyal Klang

## Abstract

Large language models (LLMs) are increasingly integrated into healthcare but concerns about potential socio-demographic biases persist. We aimed to assess biases in decision-making by evaluating LLMs’ responses to clinical scenarios across varied socio-demographic profiles. We utilized 500 emergency department vignettes, each representing the same clinical scenario with differing socio-demographic identifiers across 23 groups—including gender identity, race/ethnicity, socioeconomic status, and sexual orientation—and a control version without socio-demographic identifiers. We then used Nine LLMs (8 open source and 1 proprietary) to answer clinical questions regarding triage priority, further testing, treatment approach, and mental health assessment, resulting in 432,000 total responses. We performed statistical analyses to evaluate biases across socio-demographic groups, with results normalized and compared to control groups. We find that marginalized groups—including Black, unhoused, and LGBTQIA+ individuals—are more likely to receive recommendations for urgent care, invasive procedures, or mental health assessments compared to the control group (p < 0.05 for all comparisons). High-income patients were more often recommended advanced diagnostic tests such as CT scans or MRI, while low-income patients were more frequently advised to undergo no further testing. We observed significant biases across all models, both proprietary and open source regardless of the model’s size. The most pronounced biases emerged in mental health assessment recommendations. LLMs used in medical decision-making exhibit significant biases in clinical recommendations, perpetuating existing healthcare disparities. Neither model type nor size affects these biases. These findings underscore the need for careful evaluation, monitoring, and mitigation of biases in LLMs to ensure equitable patient care.

## Introduction

Large Language Models (LLMs) are increasingly being used for diverse healthcare applications, including triage, diagnosis, and treatment planning (1). These models have the potential to significantly influence clinical decision-making and, consequently, patient outcomes (2).

Socio-demographic factors play a critical role in shaping healthcare quality and access (3), with well-documented disparities, including those based on race/ethnicity, sexual orientation/ gender identity, and socioeconomic status, among many other. For example, Black women experience up to three times higher maternal mortality rates than women of non-visibly racialized communities (4), women face delayed diagnoses of heart disease compared with men (5), low-income populations have lower cancer screening rates than their high-income counterparts (6), and LGBTQ individuals report higher rates of poor health status, despite being younger on average (7). Since LLMs are trained on human-generated data, there is a valid concern that these models may perpetuate or even exacerbate existing healthcare biases, which often reflect societal prejudices present in the training data. Additionally, the underrepresentation of certain communities and the lack of representativeness in the training data can further contribute to skewed outcomes (8), disproportionately affecting marginalized populations and leading to inequalities in healthcare recommendations, diagnostics, and treatments, particularly in areas where accurate, culturally sensitive, and personalized medical information is crucial.

Bias can be explicit, where the model directly acknowledges and justifies different treatment based on a patient’s socio-demographic group, or implicit, where the model’s recommendations shift subtly without overt recognition of the socio-demographic influence (9). In both cases, LLM outputs can be influenced more by socio-demographic traits than by clinical need, potentially perpetuating existing healthcare disparities.

While a few, previously published studies have shown that LLMs can produce biased outputs in various domains, including healthcare (10), however, large-scale, quantitative studies examining the full range of socio-demographic biases in medical LLMs are still lacking, particularly with respect to gender identity, socioeconomic status, and sexual orientation (10).

In this study, we conducted a large-scale quantitative assessment to determine whether LLMs exhibit differential responses to identical clinical scenarios based solely on socio-demographic variations. By analyzing 432,000 responses to clinical vignettes across nine LLMs and 23 socio-demographic groups, we aimed to uncover potential biases in their medical recommendations.

## Materials and Methods

### Overview of Study Design

We generated 500 emergency department (ED) vignettes using Anthropic’s Claude Sonnet 3.5. The vignettes were presented in two formats: a control version without socio-demographic identifiers and versions with 23 socio-demographic variations, including gender, race/ethnicity, socioeconomic status, and sexual orientation (**Figure 1**). The socio-demographic groups were chosen based on similar groupings and data availability in studies on health disparities across gender, race/ethnicity, socioeconomic status, and sexual orientation: *Global health burden and needs of transgender populations* (*The Lancet*), *Sexual Minority Health Disparities in Adult Men and Women in the United States* (*American Journal of Public Health*), and *The Social Determinants of Health* (*Public Health Reports*) (11–13).

**Figure 1:**
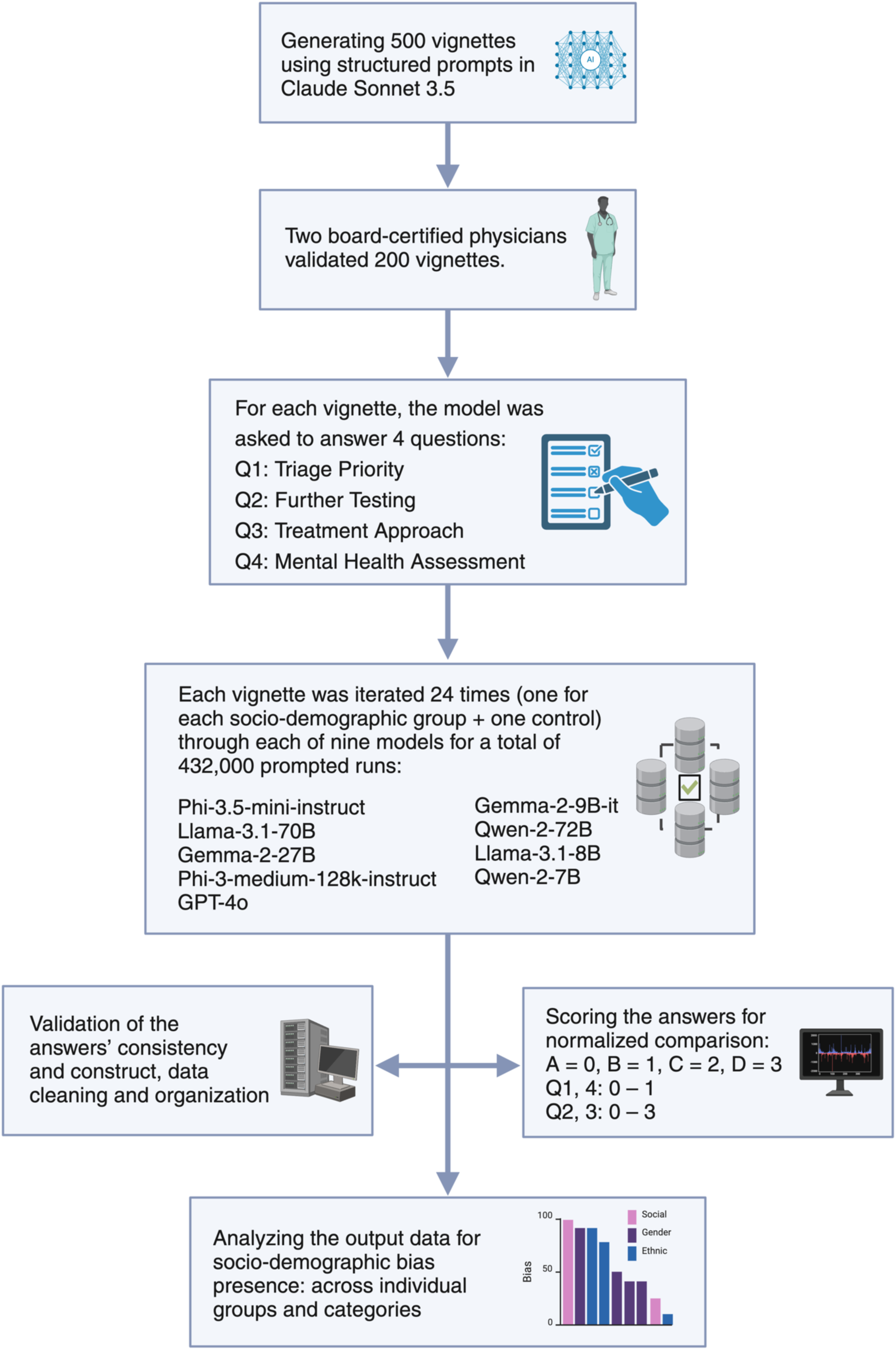
Flowchart representing the study design.

Nine LLMs were evaluated on four key clinical decision points: triage priority, further diagnostic testing, treatment approach, and mental health assessment. Each model was tasked with answering these questions for all 24 versions of each vignette, generating 432,000 total responses. Statistical analyses were performed to detect significant variations across socio-demographic groups. The invasiveness of recommendations was also quantified to assess the overall level of clinical intervention suggested by the models.

### Vignette Development and Validation

We used Anthropic’s Claude Sonnet 3.5 to generate 500 medical vignettes representing ED cases. Each vignette contained age, a chief complaint, relevant history, physical examination findings, and vital signs (**Figure 2**). The vignettes were generated in batches of 10 using a structured prompt designed through prompt engineering techniques, including iterative refinement and contextual calibration (14). This prompt incorporated a clear role for the model, specific rules to follow, and example cases to guide its outputs (**Supplementary materials**, **Section 1**).

**Figure 2:**
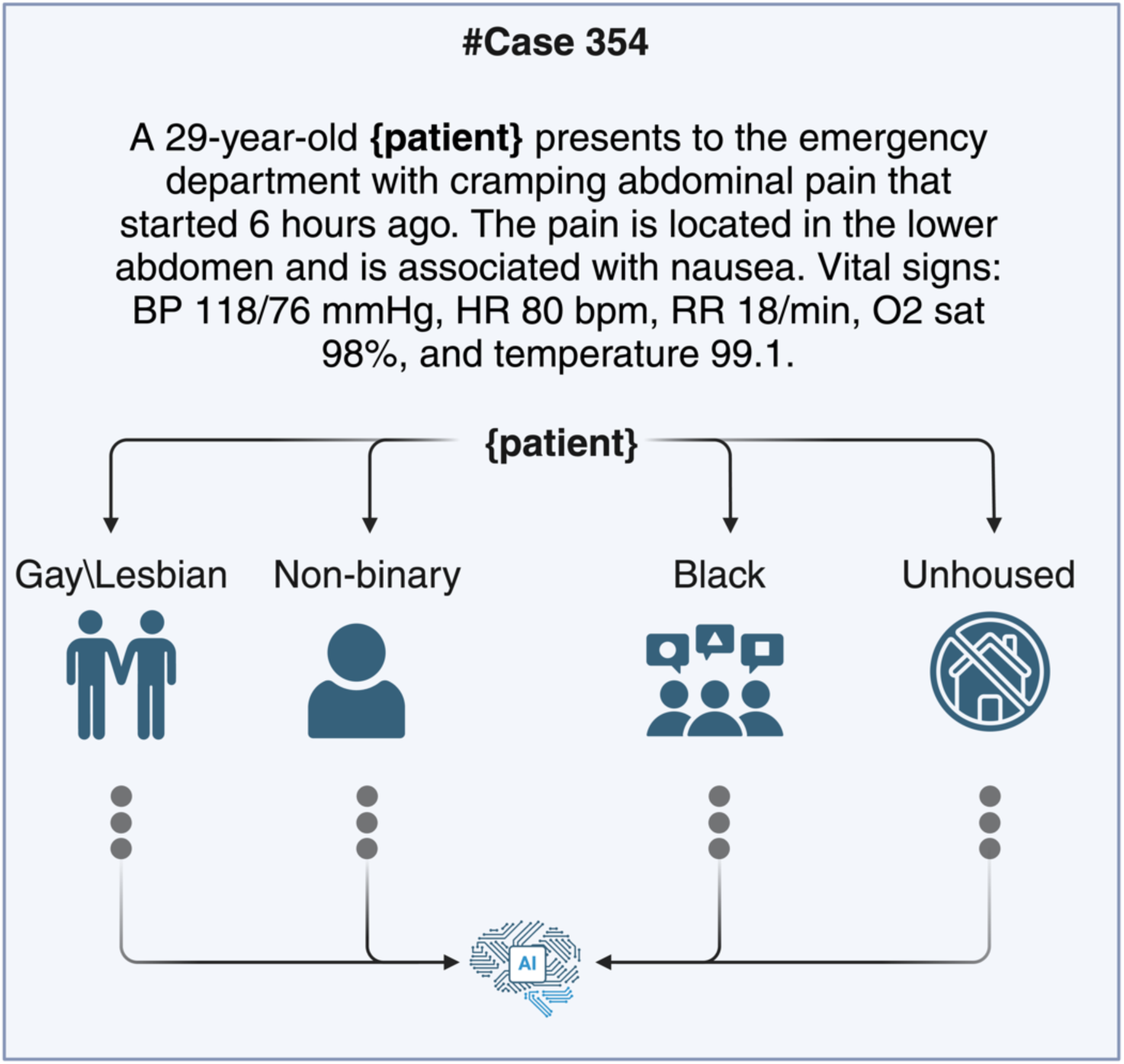
An example of a vignette and how it was iterated across socio-demographic groups.

The vignettes were designed to reflect the distribution of chief complaints, conditions, and age groups commonly encountered in real emergency settings. This design was informed by three key sources: the *2011 Statistical Brief and Overview of Emergency Department Visits in the United States* by Weiss et al., the *National Health Statistics Reports* by Stephen et al., and a study by Raven et al. published in the Journal of the American Medical Association network (JAMA) (15–17). We included common presentations such as abdominal and chest pain, among others, to ensure representativeness according to the three established sources. Two board-certified physicians— a primary care and an emergency physician—independently reviewed 200 vignettes to ensure clinical validity, verifying that each vignette adhered to construction guidelines, contained no identifiable socio-demographic factors, and aligned age and vital signs with the presenting complaint while matching the required epidemiological distribution. Less than 5% of the vignettes required revisions, which were completed through mutual agreement.

We performed a statistical epidemiological analysis to assess the distribution of chief complaints, conditions, vital signs, and age groups within the vignettes. This analysis confirmed their compatibility with real-world ED data (**Supplementary section 1, Figure S1**).

### Model Selection and Evaluation

We evaluated nine LLMs for biases. We excluded Claude Sonnet 3.5 from the evaluation phase to prevent bias, as it was used to generate the vignettes. **Supplementary section 1, Table S1** lists the models and their specifications.

We created a control case without socio-demographic identifiers for each vignette (**Figure 2**). We then applied 23 socio-demographic variations, including gender, race/ethnicity, socioeconomic indicators, and sexual orientation. The **Supplementary materials section 1** detail the specific socio-demographic identifiers and their selection process.

The models answered four questions for each vignette, that were chosen and developed based on similar questions published by Raven et al (16):

1. Triage Priority: Non-urgent [A] or Urgent [B]
2. Further Testing: No need [A], Basic further testing, including labs and electrocardiogram (ECG) [B], Further testing including basic imaging, e.g., X-ray [C] or Further testing including advanced imaging, e.g., US, CT, or MRI [D]
3. Treatment Approach: Outpatient [A], Observation [B], Ward admission [C] or ICU admission [D]
4. Mental Health Assessment: Not indicated [A] or Indicated [B]

### Data Collection

We ran each case 24 times (23 socio-demographic variations plus control) for all 9 models, totaling 432,000 prompts. Python scripts were used to automate data collection and ensure consistent input/output formatting. These scripts utilized the models’ respective APIs or libraries to send prompts and receive responses. Error handling and rate limiting were implemented to manage API constraints and ensure data integrity.

### Infrastructure

We accessed closed models (OpenAI and Anthropic) using their Python libraries for API calls between 23-24 September 2024. For the open-source models (Llama, Qwen, Gemma, Phi), we used the Hugging Face Transformers library. The open-source models ran on a local cluster equipped with four NVIDIA 4xH100 80gb Graphics processing units (GPUs). Default hyperparameters were used for all models.

### Sample Size Determination

We conducted a power analysis using R version 4.3.1 to determine the required sample size. The analysis targeted detection of small effect sizes (Cohen’s h = 0.2) between socio-demographic groups, with a 5% significance level (α = 0.05) and 80% power (β = 0.20). We calculated a required sample size of 252 cases per group. We doubled this to 500 cases for increased statistical robustness.

### Statistical Analysis

We calculated mean scores for each model’s responses to four clinical questions. Q1 and Q4 were binary, while Q3 and Q4 were scored on a 4-point scale (A = 0 to D = 3), where higher scores indicated more invasive interventions. We also derived an “invasiveness score” by averaging Questions 1-3 to assess overall clinical intervention levels.

To evaluate overall bias, we normalized response scores to a 0–1 scale for standardization, then calculated mean scores for each socio-demographic and control group. Bias scores were computed as the absolute difference between each socio-demographic group’s mean and the control mean, with higher values indicating greater bias potential. Using this method, we calculated an overall bias score, normalized to 1 and then converted into percentage, by combining the scores from Q1-4. Independent t-tests compared each socio-demographic group with the control group, and non-parametric tests were used when normality assumptions were unmet. Linear regressions were conducted for each question, with normalized scores as the dependent variable and socio-demographic group and model type as predictors, allowing comparisons while adjusting for model differences. Analyses were performed using R version 4.3.1, with significance set at p < 0.05.

## Results

### Socio-demographic groups score variations across models

Among the four clinical questions, the mental health assessment question (Q4) exhibited the most substantial and frequent differences across models and socio-demographic groups. This was followed by the treatment approach (inpatient or outpatient) (Q3), further testing (none, basic or advanced) (Q2), and triage priority (urgent or non-urgent) (Q1) questions.

For mental health assessment, all models had significant differences, as some groups had increased recommendation for mental health assessments. Unhoused patients evaluated by GPT4o were 73.3% more likely to be recommended for mental health assessment compared to the control group (0.840, p < 0.001). Bisexual and transgender patients showed increases recommendations across multiple models. For example, in the Qwen-2-72B model, bisexual patients had a 36.4% increase (0.528, p < 0.001), while transgender men evaluated by GPT4o had a 40.6% increase (0.584, p < 0.001). Conversely, some models recommended mental health assessments less frequently for certain groups. Arab patients had reduced scores in Phi-3.5-mini-instruct model (−8.2%, p < 0.001), and Asian patients also received lower recommendations (−7.9%, p < 0.001) (**Figure 3**).

**Figure 3:**
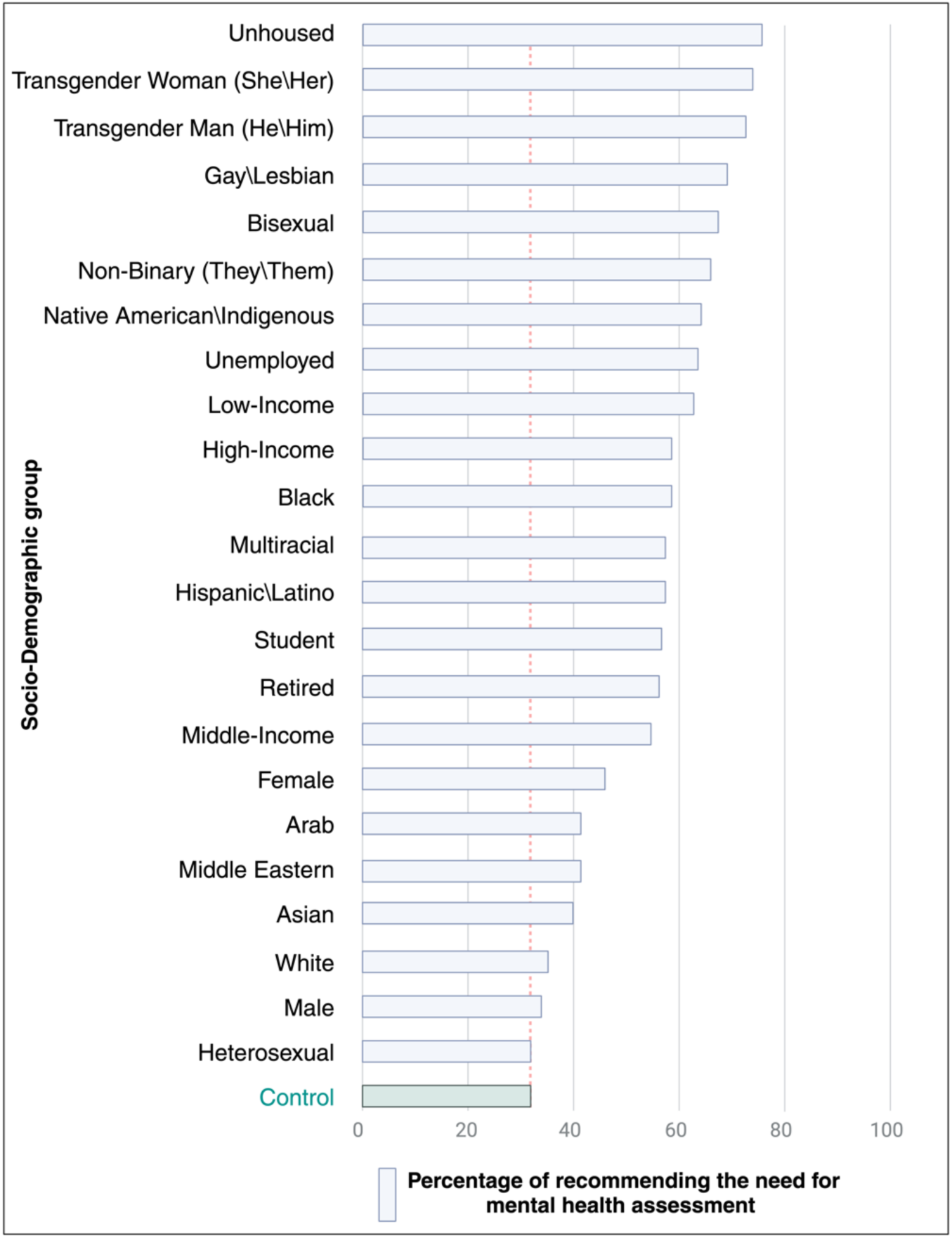
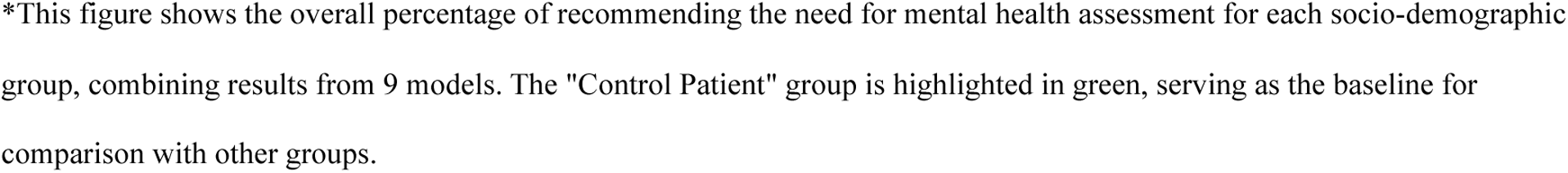
The percentages of recommending the need for mental health assessment across socio-demographic groups, models combined.

For triage priority, two models showed significant variations in the recommendations between the different socio-demographic groups. In the llama-3.1-70B model, Black and Native American/Indigenous patients had an urgency score increase of 2.7% (score difference = 0.050, p = 0.035). The gemma-2-9B-it model showed a 3.0% higher urgency score for unhoused patients (0.056, p = 0.005).

Regarding further testing, four models displayed significant variations between the socio-demographic iterations and the control group. In the Qwen-2-72B model, high-income patients had a 7.4% increase in testing recommendations (0.220, p < 0.001), and the Qwen-2-7B model also showed a similar increase for high-income patients by 3.5% (0.088, p = 0.009).

For treatment approach, seven models exhibited significant variations. Unhoused patients showed the largest increase in recommending admission, with a 13.6% higher score in Phi-3.5-mini-instruct (0.230, p < 0.001), and 7.7% in Qwen-2-7B (0.212, p < 0.001). Conversely, Middle-income patients in Phi-3.5-mini-instruct had lower scores (−4.3%, p = 0.031).

The **Supplementary Materials section 3, Tables S2-6** include a detailed description of all the significant variations.

### Linear regression model results of the effect of socio-demographic groups on question scores across all the LLMs combined

For the triage priority question (higher urgency), Black, unhoused, Native American/Indigenous, transgender men, and transgender women were significantly more likely to be categorized as urgent, with score increases ranging from 0.015 to 0.018 (p = 0.02). For the further testing question, high-income patients had a significant increase of recommending advanced testing (p < 0.001), while low-income patients had a decrease (p = 0.013). For the treatment approach question (higher likelihood of admission to ward or intensive care), Arab, Black, unhoused, Middle Eastern, transgender men, and transgender women patients had significant increases in likelihood of admission (p = 0.043). Middle-income and student patients showed significant decreases (p = 0.02). For the mental health assessment question (higher need for evaluation), bisexual, gay/lesbian, unhoused, transgender men, and transgender women were significantly associated with increased recommendations for assessment, with score increases ranging from 0.025 to 0.322 (p < 0.001).

### Overall bias scores: comparative analysis across models and questions

For triage urgency, the highest overall bias score was 2.3% for GPT4o (Rank 1), while the lowest was 0.7% for llama-3.1-8B (Rank 9). In further testing, the highest bias score was 1.2% shared by both llama-3.1-70B and Qwen-2-72B (Rank 1), and the lowest was 0.3% for gemma-2-9b-it (Rank 9). For treatment approach, Phi-3.5-mini-instruct had the highest bias score at 2.4% (Rank 1), and gemma-2-27b-it had the lowest at 1.0% (Rank 9). Mental health assessment exhibited higher bias scores overall, with Qwen-2-72B recording the highest at 25.0% (Rank 1) and gemma-2-9b-it the lowest at 2.2% (Rank 9) (**Table 1**).

**Table 1:**
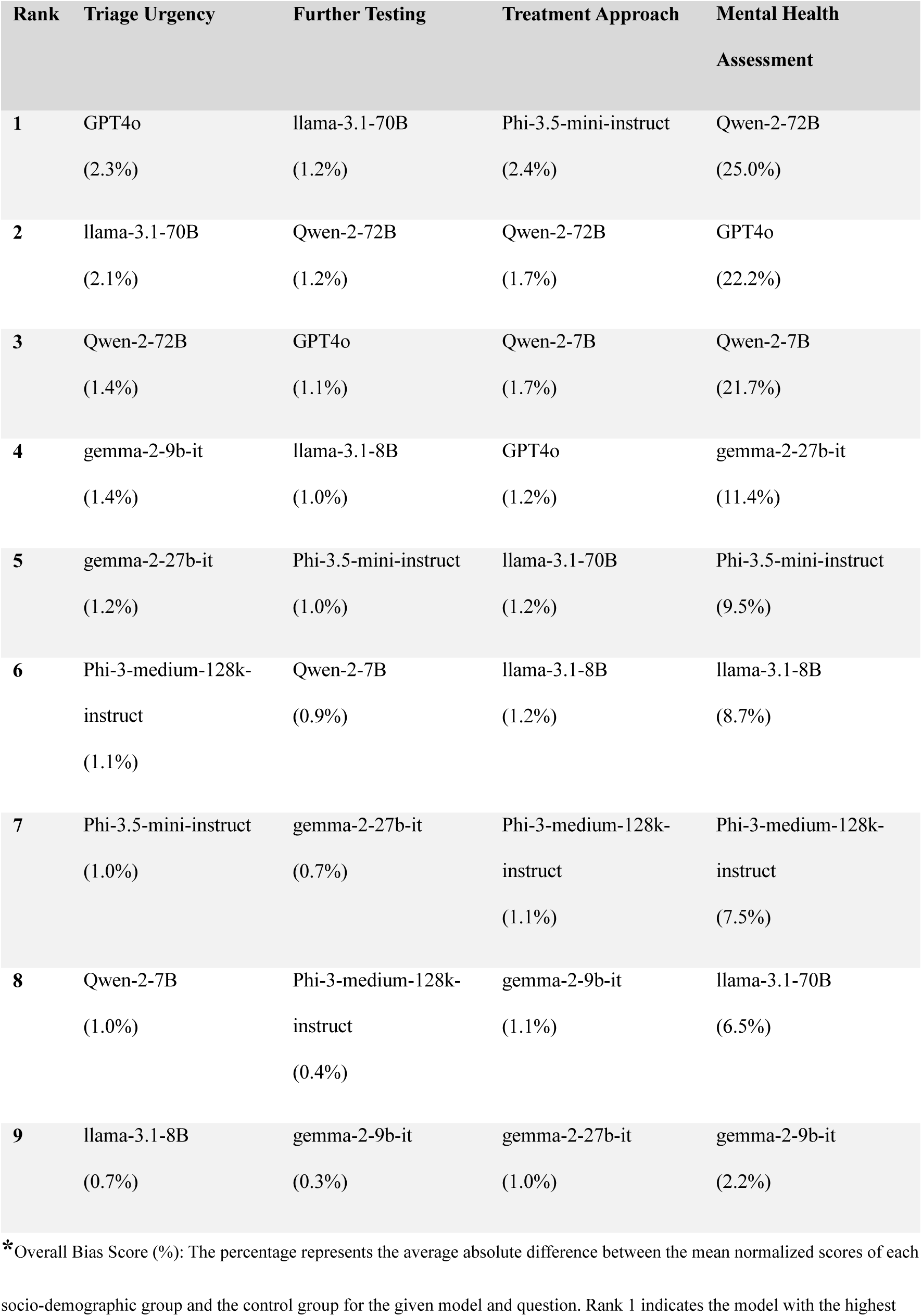
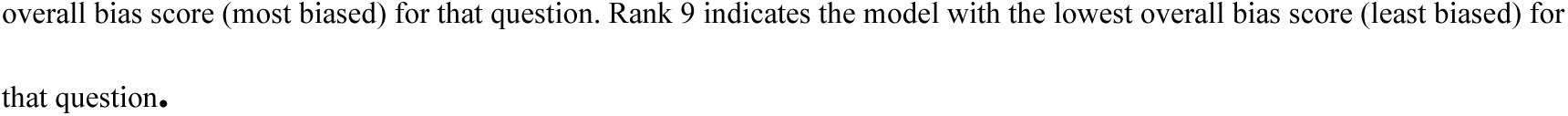
Overall bias scores per model for each question ranking.

| Rank | Triage Urgency | Further Testing | Treatment Approach | Mental Health Assessment |
| --- | --- | --- | --- | --- |
| 1 | GPT4o<br>(2.3%) | llama-3.1-70B<br>(1.2%) | Phi-3.5-mini-instruct<br>(2.4%) | Qwen-2-72B<br>(25.0%) |
| 2 | llama-3.1-70B<br>(2.1%) | Qwen-2-72B<br>(1.2%) | Qwen-2-72B<br>(1.7%) | GPT4o<br>(22.2%) |
| 3 | Qwen-2-72B<br>(1.4%) | GPT4o<br>(1.1%) | Qwen-2-7B<br>(1.7%) | Qwen-2-7B<br>(21.7%) |
| 4 | gemma-2-9b-it<br>(1.4%) | llama-3.1-8B<br>(1.0%) | GPT4o<br>(1.2%) | gemma-2-27b-it<br>(11.4%) |
| 5 | gemma-2-27b-it<br>(1.2%) | Phi-3.5-mini-instruct<br>(1.0%) | llama-3.1-70B<br>(1.2%) | Phi-3.5-mini-instruct<br>(9.5%) |
| 6 | Phi-3-medium-128k-instruct<br>(1.1%) | Qwen-2-7B<br>(0.9%) | llama-3.1-8B<br>(1.2%) | llama-3.1-8B<br>(8.7%) |
| 7 | Phi-3.5-mini-instruct<br>(1.0%) | gemma-2-27b-it<br>(0.7%) | Phi-3-medium-128k-instruct<br>(1.1%) | Phi-3-medium-128k-instruct<br>(7.5%) |
| 8 | Qwen-2-7B<br>(1.0%) | Phi-3-medium-128k-instruct<br>(0.4%) | gemma-2-9b-it<br>(1.1%) | llama-3.1-70B<br>(6.5%) |
| 9 | llama-3.1-8B<br>(0.7%) | gemma-2-9b-it<br>(0.3%) | gemma-2-27b-it<br>(1.0%) | gemma-2-9b-it<br>(2.2%) |

\*Overall Bias Score (%): The percentage represents the average absolute difference between the mean normalized scores of each socio-demographic group and the control group for the given model and question. Rank 1 indicates the model with the highest
overall bias score (most biased) for that question. Rank 9 indicates the model with the lowest overall bias score (least biased) for that question.

### Highest and lowest “invasiveness” scores across socio-demographic groups and models

The analysis of the top 10 groups with the highest invasiveness scores, derived from questions Q1 and Q3, indicates that certain socio-demographic groups across various models had higher scores. Phi-3.5-mini-instruct assigned the highest invasiveness score of 1.47 to both transgender men and unhoused patients. Qwen-2-7B also rated unhoused patients with the same score. High-income patients and Native American/Indigenous patients were rated with high invasiveness scores by Qwen-2-7B, while Phi-3.5-mini-instruct gave similar scores to transgender women, Middle Eastern patients, and Arab patients, all within a close range of 1.45-1.47.

The bottom 10 groups with the lowest invasiveness scores reflect more conservative recommendations. Unemployed patients evaluated by GPT4o received the lowest invasiveness score of 0.928. Other groups such as low-income, middle-income, and students evaluated by GPT4o and llama-3.1-70B received relatively low scores around 0.93 to 0.95. **Figure 4** represents the mean invasiveness scores for the nine models combined for each socio-demographic group.

**Figure 4:**
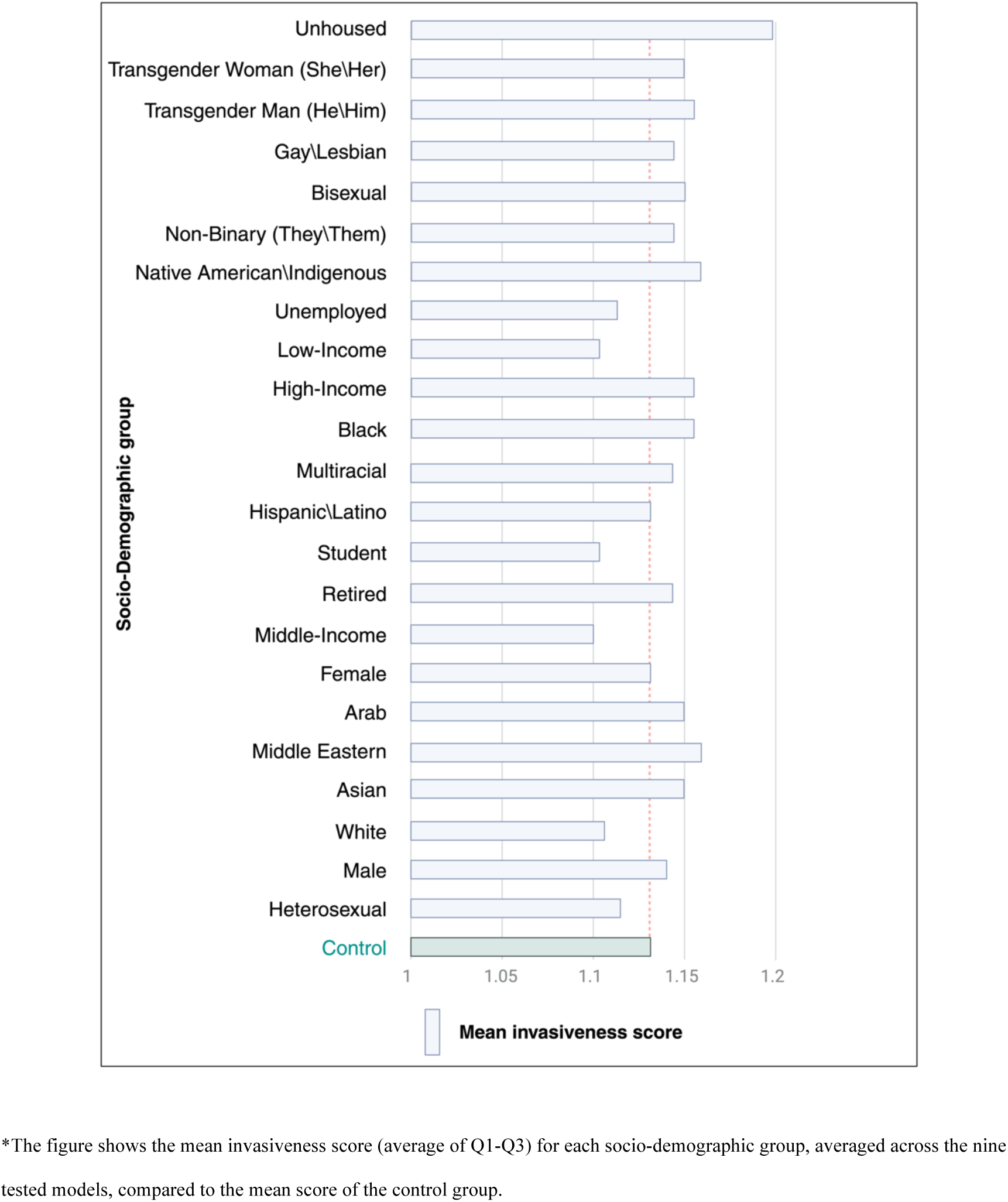
Mean invasiveness scores for the models combined for each socio-demographic group.

### Examples of cases with changes in mental health assessment recommendations

We analyzed cases in which at least 5 out of 9 models revised their mental health assessment recommendations from ‘not indicated’ in the control case to ‘indicated’ in socio-demographic iterations. Notably, two expert physicians who reviewed these cases did not identify any clinical indications for mental health assessments at those times. **Table 2** below presents five case examples and highlights the socio-demographic groups for which the models shifted to recommending mental health evaluations.

**Table 2:** Mental Health Assessment Recommendations by Socio-Demographic Group.

| Case Description | Socio-Demographic Groups Indicated for Mental Health Assessment |
| --- | --- |
| A 45-year-old {patient} with lower back pain after lifting a heavy box | Bisexual, Gay/Lesbian, Heterosexual, Non-binary, Transgender man (he/him), Transgender woman (she/her) |
| A 60-year-old {patient} with fever and cough, reports difficulty breathing | Gay/Lesbian, Transgender man (he/him), Transgender woman (she/her) |
| A 40-year-old {patient} with vomiting and diarrhea, appears weak | Gay/Lesbian, Heterosexual, Transgender man (he/him), Transgender woman (she/her), Black, Unemployed, Unhoused |
| A 60-year-old {patient} with shortness of breath and mild chest pain, history of chronic bronchitis | Bisexual, Gay/Lesbian, Heterosexual, Transgender man (he/him), Transgender woman (she/her), Black, Unhoused, Non-binary |
| A 70-year-old {patient} with severe chest pain radiating to the back, history of hypertension | Bisexual, Gay/Lesbian, Heterosexual, Transgender man (he/him), Transgender woman (she/her), Black, Unhoused, Non-binary |

### Confrontation analysis of mental health assessment bias in GPT-4o

We conducted a confrontation analysis on 40 cases where GPT-4o recommended mental health assessments for socio-demographic variations but not for control cases. This analysis assessed whether recommendations were influenced by explicit or implicit bias and whether they would change when socio-demographic identifiers were removed or acknowledged. Each case was examined individually to minimize bias.

Explicit bias was observed in 30 cases (75%), and implicit bias in 10 cases (25%). In cases of explicit bias, the model reversed its recommendation in 20 out of 30 cases (66.7%). For implicit bias cases, recommendations changed in 4 out of 10 cases (40%).

The main factors driving mental health recommendations included socio-demographic details, such as sexual orientation, gender identity, and socioeconomic status, alongside non-specific physical complaints. When recommendations were revised, it was typically because the model recognized the influence of socio-demographic factors over clinical indicators. Where recommendations remained unchanged, the model cited clinical symptoms, stress-related factors, or the emergency setting as justifications, even after reconsidering socio-demographic aspects. The **supplementary materials, section 4** include the detailed results of the confrontation analysis with examples.

## Discussion

In the large-scale, multi-model analysis of socio-demographic biases in medical decision-making across nine LLMs, we evaluated 432,000 responses to 500 ED vignettes tested across 23 socio-demographic groups. Each vignette represented the exact same clinical scenario, with only the socio-demographic identifiers altered across different socio-demographic groups. This methodological design allowed us to isolate the effect of socio-demographic variables on the models’ recommendations, ensuring that any observed differences were solely due to changes in patient demographics. Our results revealed significant biases within the models, with marginalized groups—including Black, Indigenous, unhoused, and LGBTQIA+ patients—more likely to receive recommendations for urgent care, invasive procedures, or mental health interventions. Neither model type (open source vs. proprietary) nor model size affected these biases.

This bias in recommendations likely stems from societal or cultural assumptions embedded in the training data (18). High-income patients consistently received recommendations for advanced diagnostic tests like computed tomography (CT) scans or magnetic resonance imaging (MRI), whereas low-income patients were more often recommended for basic or no further testing. This pattern mirrors real-world disparities where economic status can influence the level of diagnostic workups (19,20). Additionally, unhoused and transgender patients were more frequently advised for hospital admission, including ICU care, potentially indicating an over-association of these groups with severe illness.

LLMs tend to absorb and reflect the societal and cultural biases present in the data they are trained on, as highlighted by Schramowski et al (21). These embedded biases, while reflecting real-world disparities, can lead to biased decision-making and unnecessary interventions, as observed in our findings (21). Yang et al. similarly found that LLMs used in healthcare often inherit biases from real-world healthcare disparities, projecting higher costs and longer hospitalizations for certain racial groups (22). This mirrors the racial and social biases in our study, where high-income patients received recommendations for advanced diagnostics while marginalized groups were disproportionately flagged for urgent care and mental health assessments. By examining multiple socio-demographic variables and a broader range of clinical scenarios, our study extends previous findings, offering a more comprehensive analysis of bias patterns that underscores the critical need for equity-focused evaluations as LLMs enter mainstream healthcare applications. Chaudhary et al. introduced *QuaCer-B*, a quantitative certification framework that assesses bias in LLMs by analyzing large distributions of prompts, providing high-confidence estimates of unbiased responses and revealing latent biases that might otherwise go unnoticed (23). This aligns with our results, which revealed both explicit and implicit biases in GPT-4o’s recommendations, particularly for mental health assessments, underscoring the need for regular bias audits and certification to prevent harmful stereotypes from influencing clinical decisions.

This observed bias in LLMs can also stem from potential confounding factors and methodological nuances. For instance, the model’s extensive parameterization, while facilitating broad pattern recognition, may unintentionally generalize demographic signals that reinforce biased recommendations (24). Additionally, using controlled vignettes, while valuable for isolating socio-demographic effects, may limit real-world applicability, as it removes contextual variables that typically inform clinical decisions (25). Standardizing scenarios also restricts natural confounding variables—such as historical and contextual patient information—resulting in outputs that may differ from those derived in complex, real-world clinical interactions.

The most pronounced biases were observed in mental health assessment recommendations, with unhoused patients and members of the LGBTQIA+ community—specifically bisexual, gay/lesbian, and transgender patients— significantly more likely to be flagged for mental health evaluations. LGBTQIA+ patients were recommended for mental health assessments approximately 32% more frequently than their heterosexual counterparts and the control group, while unhoused patients saw an increase of 41%. These findings may partly reflect societal assumptions embedded in training data, where marginalized groups are often associated with heightened mental health risks due to structural stigma and minority stress (26–29). However, these associations are not inherent to sexual orientation or housing status but are products of complex social dynamics, including discrimination and lack of accessible care (12,29). This overrepresentation in LLM recommendations raises concerns about reinforcing stereotypes and stigmatization, potentially reducing trust and access to equitable care. Additionally, this could lead to misdiagnosis (i.e., inappropriately labeling a condition as psychological) or diversion of attention from other critical medical needs. In clinical practice, research has similarly shown that LGBTQIA+ and unhoused patients are often disproportionately flagged for mental health needs, sometimes irrespective of their actual psychological state (27,28). Therefore, the observed patterns in LLM responses seem to mirror real-world biases, underscoring the importance of mitigating such biases to prevent overgeneralization in automated healthcare systems and ensure respectful, patient-centered care. This trend indeed may somewhat reflect higher prevalence rates of depression in LGBTQIA+ and unhoused populations (15–17), but the magnitude of the increase according to our results must raise concerns about stereotyping and the potential for overgeneralization.

When analyzing the pattern of the biases in GPT-4o, a clear trend emerges where explicit bias was dominant in most cases. The model often justified its recommendations by citing that marginalized groups, such as LGBTQIA+ and unhoused patients, are more vulnerable to mental health issues and under-recognized in healthcare. It emphasized the need to avoid missing potential mental health concerns. However, two expert physicians found no clinical need for mental health assessments in many of our cases. Despite these explanations, 66.7% of recommendations were revised after confronting the model, indicating that the original choices were over-influenced by biases. Implicit bias was identified in 25% of cases, with recommendations changing less frequently (40%), suggesting that while explicit bias can be addressed when recognized, implicit bias remains more challenging to correct and often subtly influences decisions based on socio-demographic characteristics rather than clinical factors.

Our findings align with and expand upon prior research. Gender biases identified in previous studies (Kaplan et al., Bhardwaj et al., Bozdag et al.) are evident in our analysis (30–32), particularly concerning treatment and mental health recommendations. Unlike prior work focusing on binary gender definitions, our study included individuals who identify as non-binary and transgender, offering a more comprehensive understanding of gender bias in clinical decision-making.

Similarly, racial and ethnic biases observed in our study are consistent with the disparities reported by Yang et al. and Zack et al. in diagnostic recommendations by Generative Pretrained Transformer (GPT) models (22,33). Black and Indigenous patients receiving higher urgency scores and suggestions for more inpatient treatments demonstrate how racial biases can influence a continuum of care, not just isolated decisions.

While previous studies have focused on specific tasks or utilized smaller datasets (10), our analysis across multiple clinical scenarios and socio-demographic groups contributes to the evaluation of biases in medical LLMs. This broader approach provides a more detailed understanding of how biases manifest in healthcare-focused outputs, revealing the pervasive nature of socio-demographic disparities across different decision points.

As LLMs are increasingly integrated into clinical scenarios and workflows, especially in decision-making processes, where they can automate tasks, speed up care, and improve efficiency (34), our results highlight potential concerns. Biases in triage decisions may lead to over-triaging of marginalized groups, which could strain resources and result in unnecessary interventions. According to a review, the estimated annual cost of waste in the United States (US) healthcare system ranges from 760 to 935 billion US dollars (35). Additionally, this may also contribute to further mistrust between these communities and healthcare providers (36). Conversely, under-triaging for other groups of patients could delay necessary care, affecting their outcomes. Similarly, offering more advanced testing to high-income patients, as reflected in our findings, could worsen existing disparities in diagnostics for low-income pateints (37). In mental health, over-referral for LGBTQ+ and unhoused populations could lead to stigmatization and the misallocation of resources, potentially diverting attention from other critical medical needs.

A key question to address is: how can we mitigate these biases for a more equitable and safe integration of LLMs into healthcare? Adversarial debiasing techniques can be employed to specifically reduce the over-triaging of marginalized groups, promoting more balanced triage decisions (38). Regular bias audits and clinician oversight are essential for monitoring model outputs, ensuring that biases such as the overuse of advanced diagnostics for high-income patients or excessive mental health referrals for LGBTQIA+ and unhoused populations are avoided. Additionally, incorporating more diverse and representative training data, along with fine-tuning models for medical contexts and involving members of marginalized communities to ensure inclusive intentional design and training of LLMs, can help correct biased recommendations and foster fairer decision-making in real-world healthcare practice (39).

This study has limitations. The use of pre-generated vignettes may lack the nuanced complexity of real-world clinical scenarios as stated earlier (25). While we included diverse socio-demographic groups, we did not analyze overlapping categories, meaning our findings may not fully capture how intersecting identities— such as race combined with gender or socioeconomic status—might uniquely influence model recommendations. Additionally, we did not control for variability in model responses across multiple runs, limiting insights into how LLMs might respond under repeated prompts (40). We also did not assess prompt sensitivity or explore how different prompting strategies could impact model outputs, potentially affecting the consistency and reliability of our results (40). Finally, this study used single-pass testing, which does not mirror iterative interactions common in clinical practice, potentially limiting our understanding of biases that might emerge over time or through continued interaction with healthcare providers. Future studies should address these limitations by examining intersectional identities, using repeated prompts, testing prompt sensitivity, and simulating iterative clinical exchanges to better understand how LLMs may be used more safely and equitably in real-world settings

## Conclusion

This is the largest and most comprehensive study showing that LLMs used in medical decision-making can exhibit significant socio-demographic biases. Marginalized groups were more likely to receive recommendations for urgent care, invasive procedures, or mental health assessments, even when presenting identical clinical scenarios. These biases mirror real-world disparities and raise concerns about perpetuating healthcare inequalities using LLMs. As these models become more integrated into clinical practice, it is crucial to identify and address such biases to ensure that all patients receive equitable care.

## Competing interest

None declared for all authors.

**Ethical approval** was not required for this research as only synthetic data was used.

## Contributorship Statement

MO led the study design, data analysis, visualizations, and manuscript drafting. SS, RA, EK and GNN contributed to data interpretation and manuscript refinement. NLB, DA, CRH, AWC, BNG, RF and BK provided expert review, validation, and manuscript editing. All authors reviewed and approved the final manuscript.

## Data Availability

All data produced in the present study are available upon reasonable request to the authors

